# The Double Burden of TB/HIV co-infection: Evidence from Central Region of Ghana

**DOI:** 10.1101/2024.10.08.24315084

**Authors:** Martin Badagda Lugutuah, Daniel Boateng, Phenehance Effah Konadu, Marion Okoh-Owusu, Emmanuel Kwaku Nakua

## Abstract

The synergistic burden of Tuberculosis (TB) and Human Immunodeficiency Virus (HIV) is a significant health risk in low and middle-income economies. This study estimated the prevalence and predictors of TB/HIV co-infection in the Central region of Ghana. Using a cross-sectional study, we randomly selected five districts and employed a retrospective records review of TB cases registered between 2019 and 2022. A stepwise multivariate logistic regression was used to identify important predictors of TB/HIV co-infection. The overall prevalence of TB/HIV co-infection in the study period was 10.92%. This was highest at 55 (15.54%) among the age group 25 – 40 years. The burden of TB/HIV co-infection was highest at 36 (19.25%) in the Gomoa East district and lowest at 30 (8.38%) in Mfantseman municipal. In the adjusted model, TB patients 60 years and above had 78% lower odds of being HIV infected (AOR: 0.22; 95% CI: 0.08 – 0.66). Males were 37% less likely to be co-infected (AOR: 0.63; 95% CI: 0.42 – 0.96). Also, pulmonary-positive TB patients were 5 times more likely to be co-infected (AOR: 5.04; 95% CI: 1.71 – 14.85). Furthermore, TB patients who died in the course of treatment were 2 times more likely to be infected with HIV (AOR:2.15; 95% CI: 1.05 – 4.40). TB patients treated in the Gomoa East district were 2.7 times more likely to be HIV-infected (AOR: 2.67; 95% CI: 1.34 – 5.35). The burden of TB/HIV co-infection was moderate in the study sites and significantly associated with age, sex, bacteriological confirmation, type of TB, treatment outcome, treatment district, and year of treatment. There is a need for targeted interventions such as community awareness creation that is specific to sexually active female groups. Efforts to improve TB case detection such as health facility-based screening of patients with Respiratory Tract Infections (RTI) and contact tracing should be intensified.

## 1.0 Introduction

Globally, the “twin infection” of Tuberculosis (TB) and Human Immunodeficiency Virus (HIV) poses a major public health concern, particularly in low-resource settings [1]. TB caused by *Mycobacterium tuberculosis* is recognized worldwide as the most frequent opportunistic infection among people living with HIV [2]. In 2020, the World Health Organization (WHO) estimated TB to account for one-third of the 690,000 Acquired Immunodeficiency Syndrome (AIDS)-related deaths worldwide with approximately 84% coming from sub-Saharan Africa [3, 4]. In the 2022 WHO Global TB report, 10.6 million people were estimated to have TB in 2021 of which 6.7% of these cases were TB/HIV co-infected. This was a 4.5% increase from the 2020 estimates [5]. In countries with high HIV prevalence, living with HIV increases the likelihood of developing active TB by 26-31 times compared to HIV-seronegative individuals [3, 6, 7]. TB and HIV co-infection increases the risk of mortality due to their syndemic interaction [7, 8]. The relationship between TB and HIV is not just a simple interaction between two infectious diseases, but their combined effect accelerates adverse disease outcomes [1, 3].

Infection with both TB and HIV is known as TB/HIV co-infection. The synergistic relationship between this dual epidemic manifests in the deterioration of immunological functions which is a result of the increased level of immune activation and oxidative stress [6]. Co-infection of TB/HIV in an individual heightens the progression of each disease [9].

As in many countries of sub-Saharan Africa, Ghana is among the 41 nations accounting for the 97% global incidence of TB/HIV co-infection according to the 2015 Global TB Report [10]. Alliance and efforts from global partners have had success in lessening the effects of both infections with HIV reducing from 2.0% between 2016 and 2019 [11, 12]. Nonetheless, regional variations exist across the country. In 2017, the incidence of TB/HIV co-infection was estimated to be 21% in Ghana [10]. However, a health facility study in the Greater Accra Region estimated a co-infection prevalence of 22.9% in 2020 [13]. In another population-based study in the Volta region, TB/HIV co-infection was reported to be 19.1% in 2019 [14]. In a retrospective cohort study of 531 TB cases in two urban health facilities in Accra Ghana, treatment success was 91.2% among HIV non-infected individuals compared to 77% in HIV infected individuals [15]. In the same study, mortality associated with TB/HIV co-infection was 21.5% compared to 5.5% in those without HIV [15]. In a Nationwide survey of TB/HIV co-infection, Addo and colleagues documented an overall prevalence of 14.7% [16]. However, data from the District Health Information Management System II (DHIMS II) records Ghana’s TB/HIV co-infection as 12.9% in 2022 and 11.0% in 2023. It shows a prevalence of 7.4% and 8.4% respectively for the same period in the Central Region. With improvement in treatment regimen and public awareness, there might be a reduction in the overall prevalence of TB/HIV co-infection.

Consistent with WHO recommendations on the need for collaboration in addressing TB/HIV, TB/HIV activities have been integrated into TB and HIV services in Ghana since 2007. In this collaboration, Treatment centres are to routinely test for HIV in TB patients and vice versa. Health facilities are also required to provide a one-stop shop of services including Antiretroviral Therapy (ART) as part of the basic services for all persons diagnosed with TB/HIV co-infection [12]

Notwithstanding the glaring nature of the public health threat presented by TB/HIV synergy, there are limited epidemiological studies in the space of TB/HIV co-infection in Ghana particularly in less endowed regions such as the Central region. Previous studies in Ghana were conducted using a relatively small sample size and a limited number of sites, hence findings may not reflect that of the general population. It is therefore imperative to estimate the dual epidemic of TB and HIV regularly to track the collaborative efforts between the National TB Control Programme (NTP) and the Ghana AIDS Commission in the fight against this deadly syndemic. This will contribute to addressing program implementation bottlenecks and provide evidence for policy decisions about how best to provide high-quality integrated TB and HIV interventions at the facility and community level. From what we know, no previous studies have focused on co-infections of TB and HIV in the Central Region of Ghana. This study therefore sought to bridge this knowledge gap by determining the burden and predictors of TB/HIV co-infection in the Central region of Ghana.

## 2.0 Methods

### 2.1 Study Design

This was a cross-sectional study, involving a retrospective records review of TB cases registered between 2019 and 2022 in five districts of the Central Region of Ghana.

### 2.2 Study Setting

The Central Region occupies an area of 9,826 square kilometres, which is about 6.6% of the total land area of Ghana. The region has 22 administrative districts with the historical city of Cape Coast as the capital. About 42.1% of the region is rural with an estimated 2023 annual population of 2,989,954 with a growth rate of 2.4%[17]. TB/HIV diagnoses are mainly done in district hospital laboratories while counselling and treatment are carried out at treatment centres across districts in the region according to NTP guidelines (Central Region Annual Report, 2022). Upper Denkyira West, Mfantseman, Assin Fosu, Gomoa East, and Ajumako Enyan Essiam were randomly selected to represent the coastal, middle, and northern zones of the region as indicated in Figure 1.

**Figure 1.**
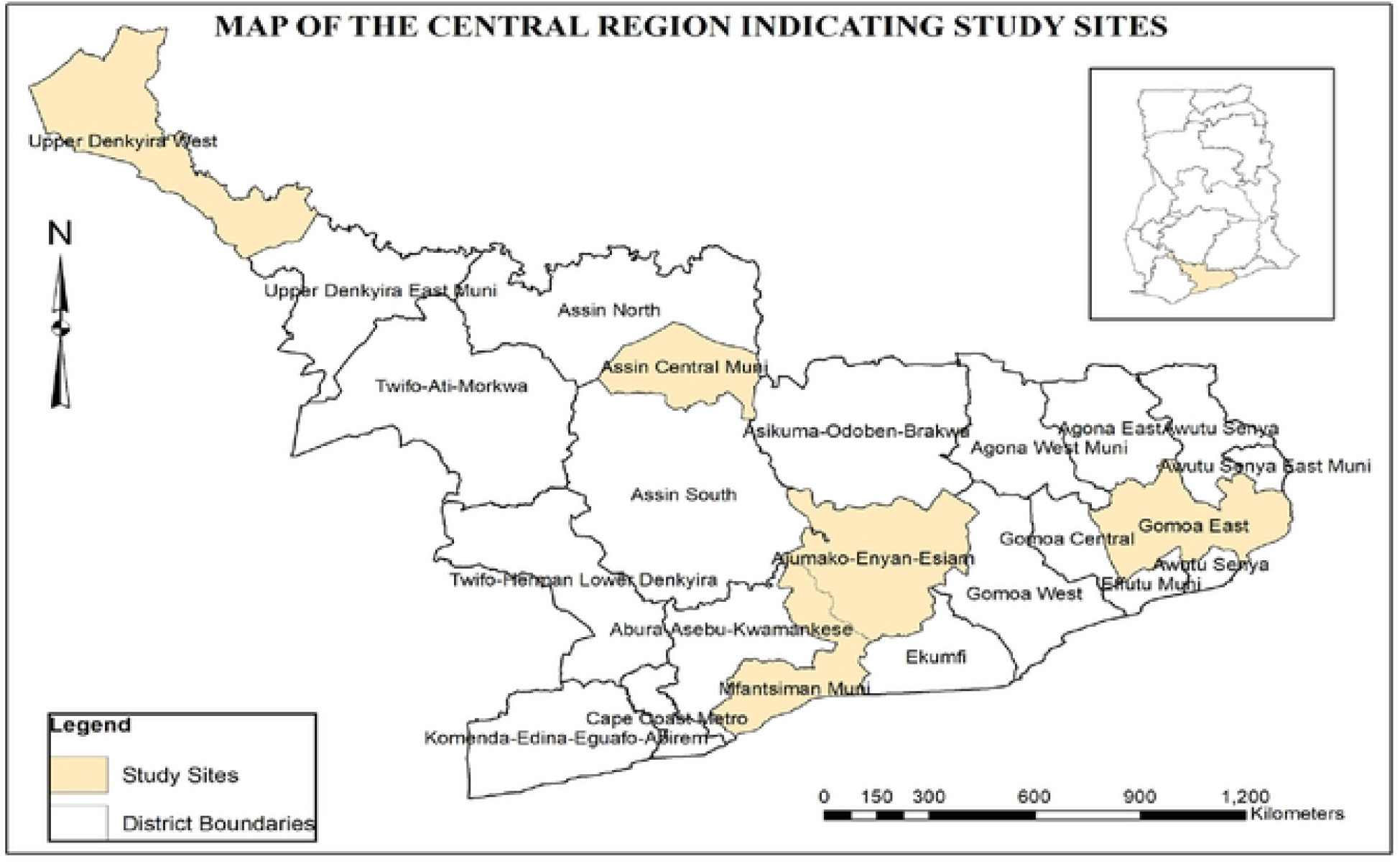
Map of Central Region of Ghana Showing Study Sites (Source: Authors construct).

**Figure 2.**
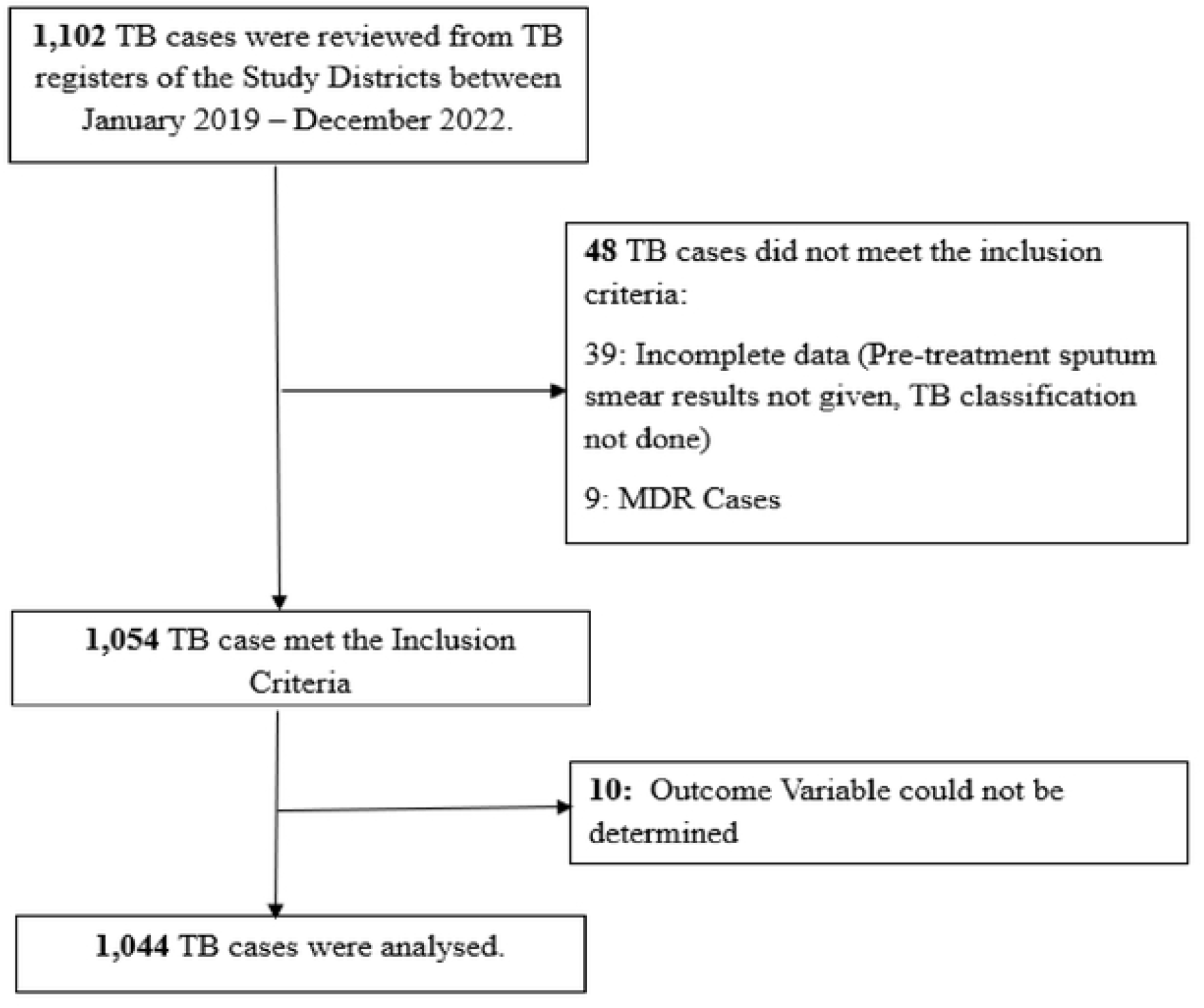
Flow Chart of sample selection based on inclusion and exclusion criteria (Source: Authors construct).

### 2.3 Sampling

Between 2019 and 2022, 1102 TB cases were found in the TB registers for the five study districts. Of these, 58 records were excluded because they were incomplete, MDR cases or the dependent variable could not be ascertained. The study population involved all persons diagnosed with TB and registered for TB care in the study districts between 1^st^ January 2019 and 31^st^ December 2022.

### 2.4 Data Collection

Data collection for this study was between 1^st^ March to 30^th^ April 2024. The data collection process involved reviewing medical records from the district TB register for information regarding age, sex, date of registration, pre-treatment bacteriological confirmation results, date of diagnosis, date of starting treatment, classification of TB, type of patient, treatment outcome and TB/HIV co-infection.

### 2.5 Study Variables and Definitions

All study variables were extracted from the district TB register. This register contains all TB cases registered in health facilities in the district. As part of the NTP guidelines TB coordinators at the district level are required to periodically update this register with TB cases reported at all health facilities in their district. The primary dependent variable for this study was TB/HIV co-infection (dichotomized as the presence or absence of TB/HIV co-infection) while treatment outcome was considered the secondary outcome (categorized as cured/treatment completed/treatment failure/lost to follow-up (defaulter) or death. Age, sex, pre-treatment bacteriological confirmation results; defined as results from biological specimens investigated with either smear microscopy, culture or GeneXpert, classification of TB and type of patient were treated as the predictor variables. All variables in this study were defined according to the WHO and National Tuberculosis Control Programme (NTP) of Ghana working definitions as presented in Table 1.

**Table 1.**
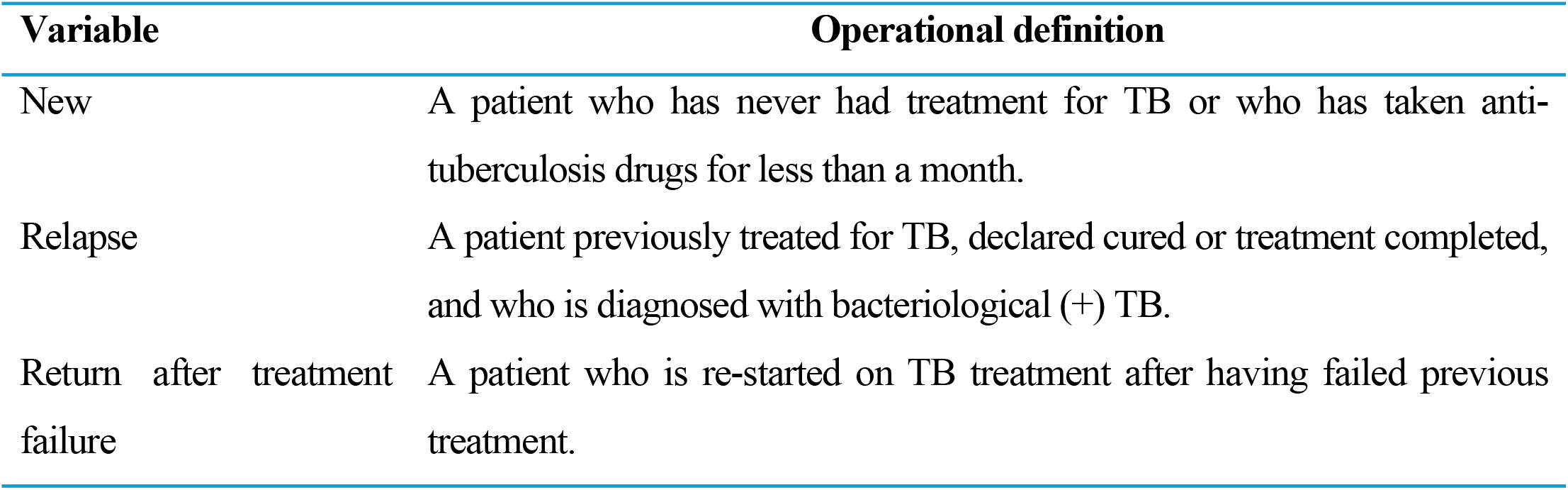
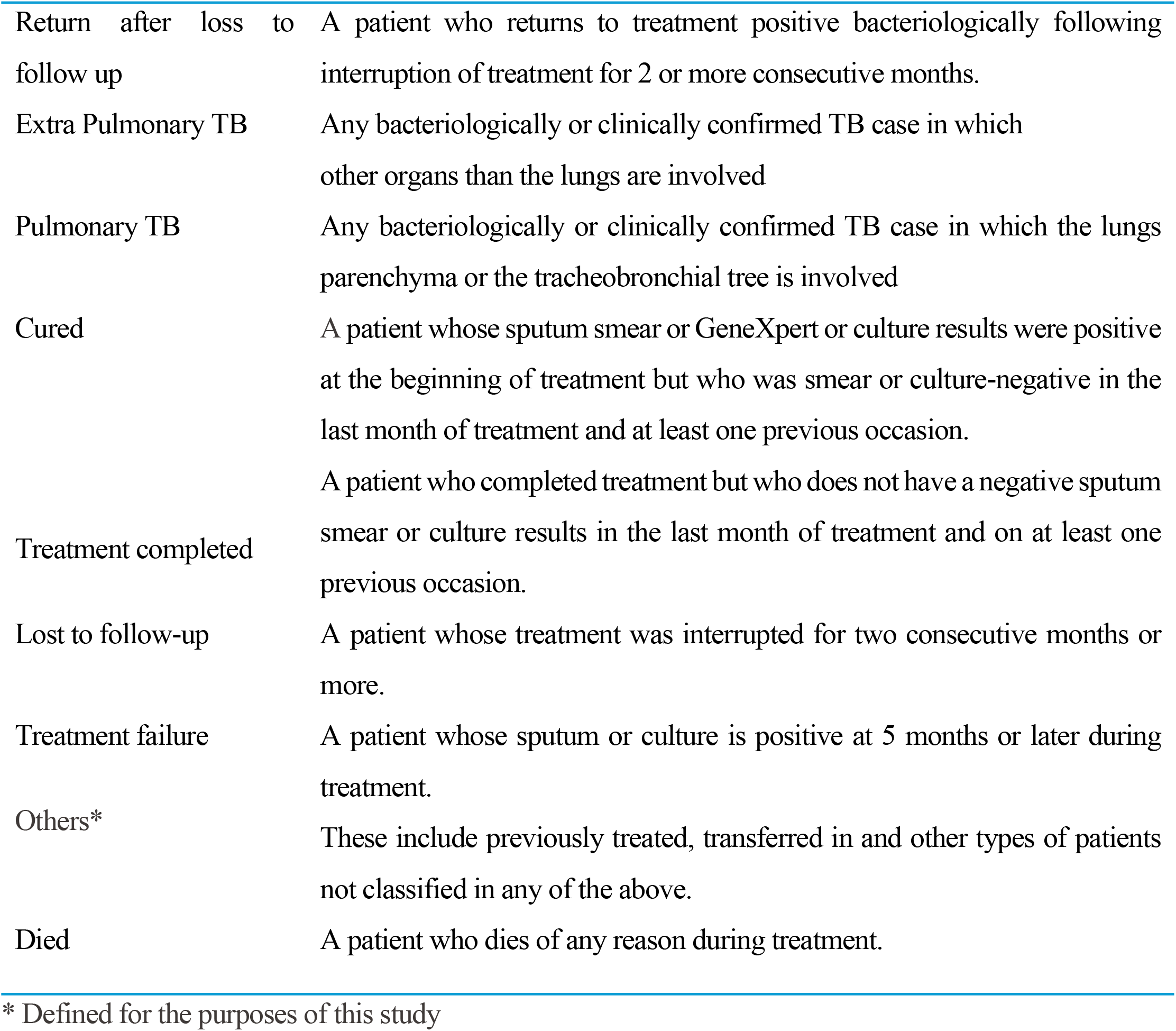
Operational definition of study variables.

| Variable | Operational definition |
| --- | --- |
| New | A patient who has never had treatment for TB or who has taken anti-tuberculosis drugs for less than a month. |
| Relapse | A patient previously treated for TB, declared cured or treatment completed, and who is diagnosed with bacteriological (+) TB. |
| Return after treatment failure | A patient who is re-started on TB treatment after having failed previous treatment. |
| Return after loss to follow up | A patient who returns to treatment positive bacteriologically following interruption of treatment for 2 or more consecutive months. |
| Extra Pulmonary TB | Any bacteriologically or clinically confirmed TB case in which other organs than the lungs are involved |
| Pulmonary TB | Any bacteriologically or clinically confirmed TB case in which the lungs parenchyma or the tracheobronchial tree is involved |
| Cured | A patient whose sputum smear or GeneXpert or culture results were positive at the beginning of treatment but who was smear or culture-negative in the last month of treatment and at least one previous occasion. |
| Treatment completed | A patient who completed treatment but who does not have a negative sputum smear or culture results in the last month of treatment and on at least one previous occasion. |
| Lost to follow-up | A patient whose treatment was interrupted for two consecutive months or more. |
| Treatment failure | A patient whose sputum or culture is positive at 5 months or later during treatment. |
| Others* | These include previously treated, transferred in and other types of patients not classified in any of the above. |
| Died | A patient who dies of any reason during treatment. |

\* Defined for the purposes of this study

**Table 2.** Demographic and Clinical Characteristics of TB Patients.

| Covariates | Frequency (n=1044) | Percentage |
| --- | --- | --- |
| **Age (years)** |  |  |
| 0 – 24 | 159 | 15.23 |
| 25 – 40 | 354 | 33.91 |
| 41 – 60 | 375 | 35.92 |
| 60+ | 156 | 14.94 |
| **Sex** |  |  |
| Female | 347 | 33.24 |
| Male | 697 | 66.76 |
| **Classification of TB** |  |  |
| Extra Pulmonary | 80 | 7.66 |
| Pulmonary – | 147 | 14.08 |
| Pulmonary + | 817 | 78.26 |
| **Type of patient** |  |  |
| New | 973 | 93.20 |
| Others | 15 | 1.14 |
| Relapse | 44 | 4.21 |
| RALTF* | 12 | 1.15 |
| **Bacteriological Confirmation** |  |  |
| Negative | 264 | 25.29 |
| Positive | 780 | 74.71 |
| **Treatment Outcome** |  |  |
| Completed | 305 | 29.21 |
| Cured | 618 | 59.20 |
| Died | 78 | 7.47 |
| LTF* | 42 | 4.02 |
| Refused Treatment | 1 | 0.10 |
| **Study site** |  |  |
| Ajumako Enyan Essiam | 226 | 21.65 |
| Assin Fosu | 154 | 14.75 |
| Gomoa East | 187 | 17.91 |
| Mfantseman | 358 | 34.29 |
| Upper Denkyira West | 119 | 11.40 |
| **Year of Diagnosis** |  |  |
| 2019 | 304 | 29.12 |
| 2020 | 217 | 20.79 |
| 2021 | 257 | 24.62 |
| 2022 | 266 | 25.48 |

RALTF\* Return After Loss to Follow-up, LTF\* Loss to Follow-up

**Table 3.** Bivariate and Multivariable analysis of TB/HIV co-infection.

| Covariates | TB/HIV<br>n=114 (%) | *p*-value<br>($\chi^2$) | COR (95% CI) | *p*-value | AOR (95% CI) | *p*-value |
| --- | --- | --- | --- | --- | --- | --- |
| **Age (years)** |  |  |  |  |  |  |
| 0 – 24 | 20 (12.58) |  | 1 |  | 1 |  |
| 25 – 40 | 55 (15.54) |  | 1.28 (0.74 – 2.22) | 0.381 | 1.31(0.73 – 2.36) | 0.369 |
| 41 – 60 | 34 (9.07) | 0.001* | 0.69 (0.39 – 1.25) | 0.220 | 0.74(0.39 – 1.39) | 0.351 |
| 60+ | 5 (3.21) |  | 0.23 (0.08 – 0.63) | 0.004* | 0.22(0.08 – 0.66) | 0.006* |
| **Sex** |  |  |  |  |  |  |
| Female | 50 (14.41) |  | 1 |  | 1 |  |
| Male | 64 (9.18) | 0.011* | 0.60 (0.40 – 0.89) | 0.011* | 0.63(0.42 – 0.96) | 0.030* |
| **Classification of TB** |  |  |  |  |  |  |
| Extra Pulmonary | 7 (8.75) |  | 1 |  | 1 |  |
| Pulmonary – | 4 (2.72) |  | 0.29 (0.08 – 1.03) | 0.055 | 0.22(0.05 – 0.85) | 0.028* |
| Pulmonary + | 103 (12.61) | 0.002* | 1.50 (0.67 – 3.36) | 0.319 | 5.04(1.71 – 14.85) | 0.003* |
| **Type of patient** |  |  |  |  |  |  |
| New | 102 (10.48) |  | 1 |  | 1 |  |
| Others | 4 (24.67) | 0.191 | 3.11 (0.97 – 9.93) | 0.056 | 2.07(0.44 – 9.58) | 0.353 |
| Relapse | 6 (13.64) |  | 1.35 (0.56 – 3.27) | 0.508 | 1.37(0.53 – 3.57) | 0.518 |
| RALTF* | 2 (16.67) |  | 1.71 (0.36 – 7.90) | 0.493 | 0.98(0.17 – 5.64) | 0.990 |
| **Bacteriological Confirmation** |  |  |  |  |  |  |
| Negative | 29 (10.98) |  | 1 |  | 1 |  |
| Positive | 85 (10.90) | 0.969 | 0.99 (0.63 – 1.55) | 0.969 | 0.31(0.14 – 0.67) | 0.003* |
| **Treatment Outcome** |  |  |  |  |  |  |
| Completed | 34 (11.15) |  | 1 |  | 1 |  |
| Cured | 60 (9.71) |  | 0.85 (0.55 – 1.34) | 0.497 | 0.80(0.49 – 1.29) | 0.359 |
| Died | 14 (17.95) | 0.238 | 1.74 (1.88 – 3.44) | 0.010* | 2.15(1.05 – 4.40) | 0.036* |
| LTF* | 6 (14.29) |  | 1.33 (0.54 – 3.38) | 0.552 | 1.20(0.45 – 3.19) | 0.713 |
| Refused Treatment | 0 |  |  |  |  |  |
| **Study site** |  |  |  |  |  |  |
| Ajumako Enyan Essiam | 20 (8.85) |  | 1 |  | 1 |  |
| Assin Fosu | 18 (11.69) |  | 1.36 (0.70 – 2.67) | 0.367 | 1.53(0.72 – 3.28) | 0.656 |
| Gomoa East | 36 (19.25) | 0.001* | 2.46 (1.37 – 4.41) | 0.003* | 2.68(1.34 – 5.35) | 0.005* |
| Mfantseman | 30 (8.38) |  | 0.94 (0.52 – 1.70) | 0.843 | 0.95(0.48 – 1.89) | 0.888 |
| Upper Denkyira West | 10 (8.40) |  | 0.94 (0.43 – 2.09) | 0.889 | 1.58(0.66 – 3.82) | 0.304 |
| **Year of Diagnosis** |  |  |  |  |  |  |
| 2019 | 44 (14.47) |  | 1 |  | 1 |  |
| 2020 | 16 (7.37) |  | 0.47 (0.26 – 0.86) | 0.014* | 0.51(0.28 – 0.95) | 0.034* |
| 2021 | 30 (11.67) | 0.048 | 0.78 (0.47 – 1.28) | 0.330 | 0.90(0.54 – 1.52) | 0.704 |
| 2022 | 24 (9.02) |  | 0.59 (0.35 – 0.99) | 0.047 | 0.73(0.42 – 1.29) | 0.282 |
| **Overall Prevalence** | **114 (10.92)** |  |  |  |  |  |

\*Statistical significance, RALTF\* Return After Loss to Follow-up, COR Crude Odds Ratio, AOR Adjusted Odds Ratio.

### 2.6 Data Management and Statistical Analysis

Data from the TB registers was extracted into a Microsoft Excel 365 spreadsheet for wrangling before being exported to STATA version 17 for analysis. Graduate National Service Personnel were trained as research assistants to extract data. Variables such as Age, sex, pre-treatment bacteriologically confirmation results, classification of TB and type of patient, treatment outcome and TB/HIV co-infection were extracted from the TB register. Consistency and completeness checks were applied to ensure the data extracted was fit for analysis. Descriptive statistics were reported using frequency, percentages, tables, and graphs. To estimate the burden of the primary dependent variable, the total number of TB patients testing positive for HIV was used as the numerator against the total number of TB patients testing for HIV over the study period. A stepwise multivariate logistic regression was used to identify important predictors of TB/HIV co-infection in the study area. At a 95% confidence interval, odds ratios (OR) were used to assess the association between the explanatory and outcome variables. A *p*-value of <0.05 was considered statistically significant.

### 2.7 Ethical Considerations

Ethical approval was obtained from the Ethics Review Committee of the Ghana Health Service **(Protocol Number: GHS-ERC 016/02/24)**. Written permission was obtained from the Central Regional Health Directorate. The PI also signed a confidentiality statement with the five study District Health Directorates.

## 3.0 Results

### 3.1 Demographic and Clinical Characteristics of TB Patients

There were 1,044 study patients in this study. A majority (66.8%) were males, and the mean age was 42 ±17 years. Eight Hundred and Seventeen (78.3%) of the cases were Pulmonary Positive while 80 (7.7%) were Extra Pulmonary. For the category of TB cases, 973 (93.2%) were new TB cases, 44 (4.2%) were relapse cases, and 12 (1.2%) were returned after loss to follow–up. Concerning treatment outcome, 618 (59.2%) were cured, 305 (29.2%) completed treatment, and 78 (7.5%) died. Of the total 1044 TB cases, 358 (34.3%) were recorded in Mfanteman district, 226 (21.7%) in Ajumako Enyan Essiam District, 187 (17.9%), 154 (14.8%) and 119 (11.4%) for Gomoa East, Assin Fosu and Upper Denkyira West districts respectively. With regards to years of treatment, majority 304 (29.12%) of the cases were treated in 2019 while 217 (20.79%) were treated in 2020.

### 3.2 Burden of TB/HIV Co-infection

The overall prevalence of TB/HIV co-infection in the study period was 10.92%. This was highest 55 (15.54%) among age group 25 – 40 years compared to 5 (3.21%) in patients 60 years and above. In 2019, 44 (14.47%) TB patients were infected with HIV. This dipped significantly to 16 (7.37%) in 2020 and subsequently rose to 30 (11.67%) in 2021, however, in 2022, it decreased to 24 (9.02%). Patients aged 25 – 40 years had higher odds (OR:1.28; 95% CI: 0.74 – 2.22; *p* = 0.381) of being co-infected compared to those aged ≤ 24 years, although this was not statistically significant. In this study, TB patients 60 years and above had significantly lower odds (OR: 0.23; 95% CI: 0.08 – 0.63; *p* < 0.05) of being co-infected compared to ≤ 24 years. Concerning sex, prevalence was 50 (14.41%) among females compared to 64 (9.18%) males. Males had significantly lower odds (OR: 0.60; 95% CI: 0.40 – 0.89; *p* < 0.05) of being TB/HIV co-infection. With a prevalence of 103 (12.61%), pulmonary-positive TB patients were twice as likely to be co-infected compared to extrapulmonary patients. This was however not statistically significant. We also found that 14 (17.95%) of patients who died in the course of treatment were 1.7 times more (OR: 1.72; 95% CI: 1.88 – 3.44; *p* < 0.05) likely to be co-infected compared to those who completed treatment. The burden of TB/HIV co-infection was highest at 36 (19.25%) in Gomoa East and lowest at 30 (8.38%) in Mfantseman as shown in Figure 3. Additionally, compared to TB patients treated in the Ajumako Enyan Essiam district, those in the Gomoa East district were 2.5 times more likely to be co-infected with HIV (OR: 2.46; 95% CI:1.37 – 4.41; *p* = 0.003). The prevalence of TB/HIV co-infection has been unstable during the four-year study period. Our study showed that TB patients treated in 2020 were 53% less likely to be HIV positive (OR: 0.47; 95% CI: 0.26--0.86, *p* = 0.014) compared to their counterparts in 2019.

**Figure 3.**
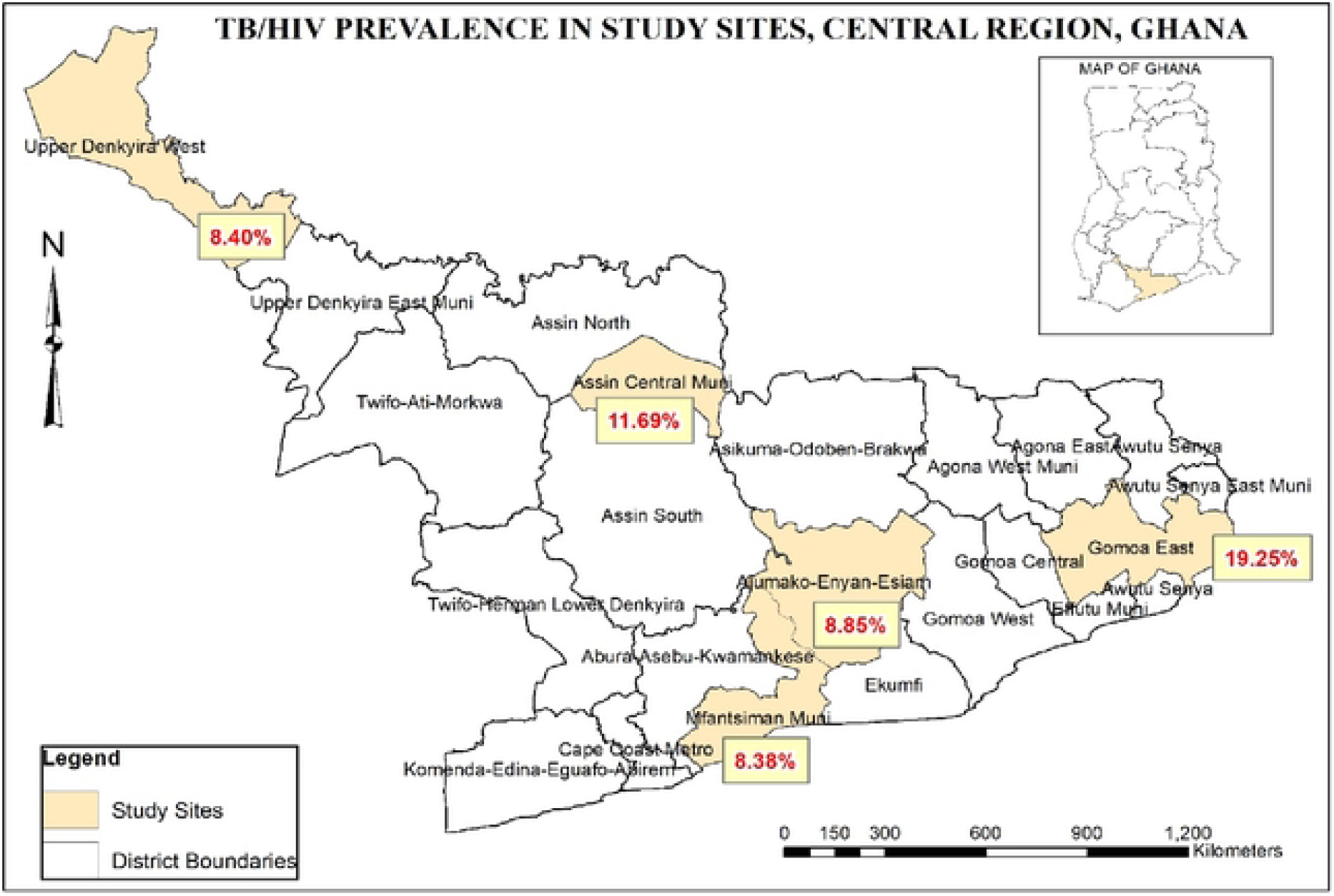
TB/HIV Prevalence in Study Sites (Source: Authors construct).

### 3.3 Predictors of TB/HIV Co-infection

In the adjusted model, TB patients 60 years and above had 78% lower odds of being HIV infected compared to patients who were ≤ 24 years of age (AOR: 0.22; 95% CI: 0.08 – 0.66; *p* = 0.006). With regards to sex, males were 37% less likely to be co-infected compared to females (AOR: 0.63; 95% CI: 0.42 – 0.96; *p* = 0.030). Also, pulmonary-positive TB patients were 5 times more likely to be co-infected compared with Extra pulmonary patients (AOR: 5.04; 95% CI: 1.71 – 14.85; *p* = 0.03). Pulmonary-negative cases however had a 78% lower odds of being co-infected (AOR: 0.22; 95% CI: 0.05 – 0.85; *p* = 0.028). Additionally, bacteriologically confirmed TB cases had almost 70% lower odds of being co-infected compared with those clinically confirmed (AOR: 0.31; 95% CI: 0.14 – 0.67; *p* = 0.003). Furthermore, TB patients who died in the course of treatment were 2 times more likely to be infected with HIV compared with those who completed treatment (AOR:2.15; 95% CI: 1.05 – 4.40; *p* = 0.036). Loss to follow-up TB patients also had a 20% higher odds of being HIV co-infected, this was however not statistically significant. TB patients treated in the Gomoa East district were 2.7 times more likely to be HIV infected compared to those treated in the Ajumako Enyan Essiam district (AOR: 2.67; 95% CI: 1.34 – 5.35; *p* = 0.005). TB patients treated in the year 2020 had a 49% lower odds of being HIV positive compared to those registered in 2019 (AOR: 0.51; 95% CI: 0.28 – 0.95; *p* = 0.034). In this study, we observed that age, sex, treatment outcome, treatment district, and year of treatment were significantly associated with TB/HIV co-infection.

## 4.0 Discussion

The HIV epidemic has considerably influenced the epidemiology of TB [18]. Our study examined the dual burden of TB/HIV co-infections in the Central Region of Ghana between 2019 and 2022. The prevalence of TB/HIV co-infection in the study area was 10.92%, higher than WHO 2022 global estimates of 6.7% [19]. This corroborates with studies conducted in Nepal [9], Ghana [16, 19] and Ethiopia [21]. Our findings are however lower than the findings of similar studies conducted in Ghana (21.5%), (18%), (18.6%) [1,11,15] and Kazakhstan (22 %) [22]. The current findings could be the results of efforts by the Ghana Health Service to reduce the TB/HIV burden through improvements in treatment regimens.

In the present study, TB patients aged ≤ 24 years were at a higher risk of HIV infection compared to those aged 60 years and above. We found that TB patients aged 60 years and above had 78% lower odds of HIV infection. This finding is in congruence with [23]. These findings are considerably different from what has been reported by other studies in Ghana [11] and northeast Ethiopia [18]. The difference in findings could be attributed to the difference in sample selection where the other studies exclude TB patients below 18 years. Although men are at a greater risk of TB and consistent with findings from several epidemiological studies [5, 11, 12, 23, 24], we found males were 37% less likely to be co-infected with HIV. This is explained by the biological make-up of females which makes them more susceptible to the risk of acquiring HIV infection [26].

Findings from our study suggest that co-infection was 5 times more likely in pulmonary-positive TB patients compared with extrapulmonary patients. This is similar to results reported in Ghana [15]. However, these findings are incongruence with what was observed by [27] in Harare, Zimbabwe. A study conducted in Ethiopia found no statistical difference between type of TB and HIV co-infection [23]. The limited capacity to diagnose extrapulmonary TB in the study settings could account for this disparity.

This study showed that almost bacteriologically confirmed TB cases had a 70% lower odds of co-infection compared with clinically diagnosed TB. Limited studies have considered this type of analysis. It is more likely that bacteriologically confirmed TB patients would take TB treatment and counselling more seriously because the results have been confirmed in a laboratory compared to those with bacteriologically negative results who may have some doubts.

In this study, the odds of HIV infection among TB patients who died in the course of treatment was 2.17 times compared with those who completed treatment. These findings are analogous to those found in similar settings of the same region [28] and in Malaysia [29]. According to the current literature on TB/HIV co-infection, these two duos in an individual increase the risk of mortality due to their syndemic interaction [1, 3, 9, 19].

There was a disproportionate rate of TB/HIV co-infection in the study area ranging from 8.35% to 19.25%. Our study demonstrated that TB patients who live in the Gomoa East treatment site were 2.4 times more likely to be co-infected with HIV compared to those who lived in the Ajumako Enyan Essiam district. Limited studies have been conducted in the study sites. However, the district TB coordinator cited the existence of the erstwhile Budumburam camp as a contributing factor as the area which is located in the district is known to be a highly endemic of HIV. Prevalence of TB/HIV co-infection in the WHO AFRO Region shows a range of 2.9% to 72.3%, with a pooled prevalence of 23.5% [13]. The finding from this study is also lower than and range of 10.8% to 26% documented in the Volta region of Ghana [12]. These differences could be attributed to the fact that TB/HIV co-infections have been reducing globally since 2008 [30].

TB patients diagnosed in 2020 in this study were 49% less likely to be co-infected with HIV compared to those diagnosed in 2019. We found a low (20.79%) TB case detection in all the study sites in 2020. The WHO Global Tuberculosis Report 2022 has stated the negative impact of the COVID-19 pandemic on TB management [19]. Osei and colleagues in their study contended that the COVID-19 pandemic resulted in an abrupt decline in TB notification and HIV testing by 20.5% and 40.3% respectively [31]. With reduced cases of TB detected in 2020, there is a high likelihood of a corresponding low detection of TB/HIV co-infection as TB cases are tested for HIV after TB detection through the TB and HIV collaboration.

This was a cross-sectional study, based on a retrospective records review of existing health facility TB records. We could not independently verify the HIV and TB status of patients. Also, other important variables such as education, occupation, nutrition, alcohol use, and smoking were not accessed in this study. Notwithstanding these limitations, the study generated valuable evidence into the prevalence and predictors of HIV among TB patients in the Central region of Ghana. These findings may guide TB and HIV control program managers and policymakers to improve the implementation of TB/HIV integration activities to reduce this dual epidemic.

## 5.0 Conclusion

The burden of TB/HIV co-infection was moderate in the study sites and significantly associated with age, sex, bacteriological confirmation, type of TB, treatment outcome, treatment district, and year of treatment. There is a need for targeted interventions such as community awareness creation that is specific to sexually active female groups, especially in the Gomoa East District. Efforts to improve TB case detection such as health facility-based screening of patients with Respiratory Tract Infections (RTI) and contact tracing should be intensified. Logistics constraints to the testing for HIV in TB patients should also addressed by program managers to improve the detection of co-infections. Future prospective studies of TB/HIV co-infection should be conducted in the study settings to understand the risk of co-infection properly.

## Data Availability

Data will be made available by the corresponding Author on request

## Authors Contributions

**Conceptualization:** Martin Badagda Lugutuah

**Data curation:** Martin Badagda Lugutuah, Phenehance Effah Konadu

**Formal Analysis:** Martin Badagda Lugutuah, Daniel Boateng

**Validation:** Daniel Boateng, Emmanuel Kwaku Nakua

**Writing Original draft:** Martin Badagda Lugutuah, Daniel Boateng, Phenehance Effah Konadu

**Review and Editing:** Daniel Boateng, Marion Okoh-Owusu and Emmanuel Kwaku Nakua

## Supporting Information

The correspondence author will make other information available upon request.

## Acknowledgements

We acknowledge the support of the Central Regional Health Directorate and Heads of selected study sites.

